# TRAVEL-ASSOCIATED CHIKUNGUNYA VIRUS INFECTION, MEXICO, 2025

**DOI:** 10.64898/2026.01.07.26343440

**Authors:** Daniel Canul Canul, K. Jacqueline Ciau-Carrillo, J. Reyes Canche Pech, Cesar Cañas-Alamilla, Jesús Kú-Cachón, Ana Osorio-Medrano, Maria E. Lopez-Novelo, Igrid García González, James Earnest, Daniel Limonta, Norma Pavia-Ruz, Pablo Manrique-Saide, Fabián Correa-Morales, Guadalupe Ayora Talavera, Laura Conde-Ferraez, Azael Che-Mendoza, Jorge Palacio-Vargas, Rafael Valdez-Vázquez, Carlois Frederico Campelo de Albuquerque e Melo, Gonzalo Vazquez-Prokopec, The Arbovirus Co-Circulation (ARCC) Research Consortium, Tetyana I. Vasylyeva, Marina Escalera-Zamudio, Miguel A. Garcia-Knight, Henry Puerta-Guardo

**Affiliations:** Universidad Autónoma de Yucatán, Centro de Investigaciones Regionales Dr. Hideyo Noguchi, Merida, Yucatan, Mexico; Biomédicos de Mérida, Laboratorios Clínicos, Mérida, Yucatan, México; University of Central Florida, Burnett School of Biomedical Sciences, Orlando, Florida, USA; Universidad Autónoma de Yucatán, Unidad Colaborativa de Bioensayos Entomológicos (UCBE), Merida; Secretaría de Salud México, Centro Nacional de Prevención y Control de Enfermedades (CENAPRECE), Mexico City, Mexico; Pan American Health Organization; Department of Environmental Sciences, Emory Universiry; University of California, Irvine, Joe C. Wen School of Population and Public Health, California, USA; Genetics Institute, University College London, London, UK; Instituto de Investigaciones Biomédicas, Universidad Nacional Autónoma de México, Mexico City, Mexico; Wellcome Sanger Institute, Hinxton, United Kingdom

**Author notes:** Address for correspondence: Henry Puerta-Guardo, Laboratorio de Virología, Centro de Investigaciones Regionales Dr. Hideyo Noguchi, Universidad Autónoma de Yucatán, Calle 96 s/n x Av. Jacinto Canek y Calle 47, Paseo de las Fuentes, CP 97225, Merida, Yucatan, Mexico;. These authors contributed equally to this work (Daniel Canul Canul, K. Jacqueline Ciau-Carrillo). Additional members of the ARCC Research Consortium are listed at the end of this article. These senior authors contributed equally to this article (Marina Escalera-Zamudio, Miguel A. Garcia-Knight, Henry Puerta-Guardo). Additional members of the ARCC Research Consortium: Julius Lutwama and John Kayiwa (UVRI, Uganda), (UVRI, Uganda), Tierra Smiley and David J Wolking (UC Berkeley, USA) and Benard Sebidde (Gorilla Doctors, Uganda).

**Keywords:** Chikungunya virus, phylogenetics, arbovirus, Mexico

## Abstract

In November 2025, a traveler from Cuba tested positive for chikungunya virus upon arrival to Mexico. The virus belonged to the East-Central-South-African lineage, clustering with a clade prevalent in Brazil. Ten years after the last chikungunya epidemic in Mexico, strengthened surveillance is required to anticipate transmission of this emergent lineage.

## Introduction

Chikungunya (CHIKV; genus *Alphavirus*, family *Togaviridae*) is a re-emerging *Aedes* borne virus that causes an acute febrile illness (incubation period typically 3-7 days) which is generally self-limiting, but can produce severe polyarthralgia, with a chronicity rate of 44% and, in rare cases, neurologic complications and death. Globally, around 35 million CHIKV infections occur each year, including large epidemics across tropical and subtropical regions, with the Americas being particularly affected (up to 3.7 million reported cases) (*1*). In areas endemic for dengue and Zika viruses, symptoms such as fever, rash, malaise and joint pain, may overlap amongst the three infections, highlighting the importance of accurate diagnostics and genomic surveillance (*2*).

Three major CHIKV lineages are currently recognized: the West African (WA), Asian (AS), and East-Central-South-African (ECSA). Whilst the WA lineage is associated with enzootic transmission and local outbreaks in Western Africa, the ECSA and Asian lineages have widely expanded into new regions (*2*). In 2013, CHIKV affected 50 countries and territories in the Americas and led to explosive epidemics that subsequently subsided, particularly those driven by the Asian lineage with *Aedes aegypti* as the primary vector (3). Nevertheless, resurgent epidemics have occurred, mainly in South America, and in 2025, more than 110,000 confirmed cases have been reported in the Region of the Americas, largely from Brazil, with >38,000 suspected cases reported in Cuba (4). Despite intense transmission, regional genomic surveillance remains uneven, with most genomes generated in Brazil and none publicly available from Cuba (5).

As of December 3^rd^, 2025, Mexico reported four chikungunya fever (CHIKF) cases, with unclear infection histories, in the southern states of Quintana Roo (three) and in Yucatan (one) (*6*). Yucatan is a recognized hotspot for Aedes-borne diseases (*7*), with serologic evidence of recent chikungunya virus exposure (IgG and IgM CHIKV seroreactivity of 24% and 2.6%, respectively), in children aged 2–15 years residing in urban areas in the state (*8*), suggestive of possible transmission.

### The Study

A healthy adult male traveled from the USA to Cuba in late October, 2025, and four days later flew to Merida, Yucatan, Mexico. Two days upon arriving in Mexico, he developed mild fever (<38 °C), myalgia, arthralgia, malaise, and cold-like symptoms (Figure 1a). The following day, the patient reported pruritus, as well as a maculopapular rash on the posterior neck and abdomen (Figure 1b). CHIKV infection was confirmed by RT-qPCR (Ct 20.92) seven days after the patient had arrived in Cuba. Self-reporting by the patient indicated that the rashes on the neck and abdomen persisted for over two weeks and one month, respectively. Informed consent was obtained, and identifying information was omitted to ensure the patient’s privacy and confidentiality. A viral isolate was obtained from plasma after inoculation onto Vero cell monolayers and the observation of cytopathic effect after 48hrs (Ct of isolate: 14.59) (Figure 1c).

**Figure 1.**
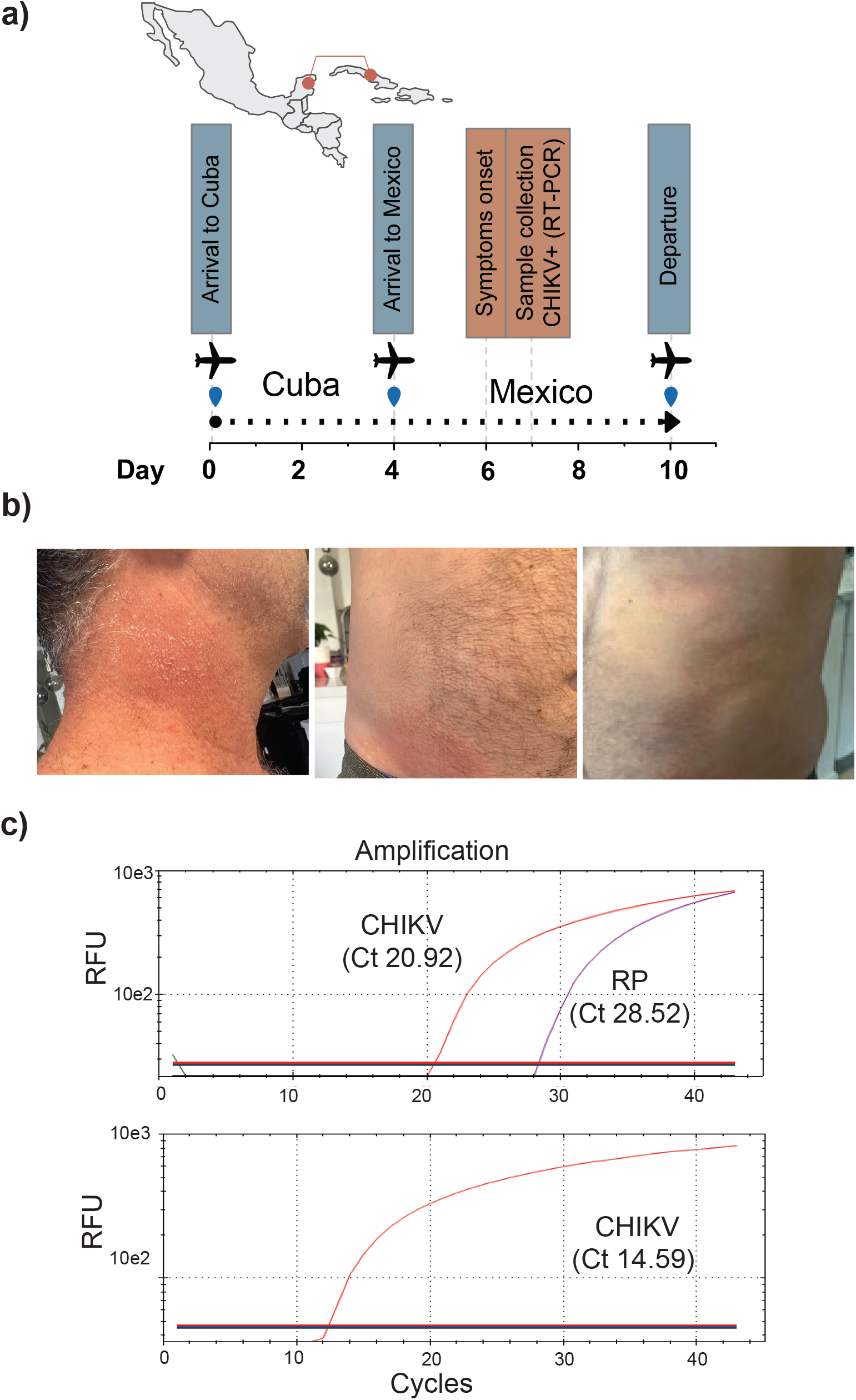
Travel-associated CHIKV case confirmation by detection of CHIKV RNA from plasma and a cell culture isolate. **(a)** Timeline for travel-associated CHIKV case diagnosis and symptom development. **(b)** Maculopapular rash on the posterior neck and left and right side of the abdomen Rash **(c)** RT-qPCR amplification plots for the detection of CHIKV RNA from plasma (top panel), and from cell supernatant derived from Vero-infected cells (lower panel). The RNase P (RP) human gene was used as reference control for amplification. RFU, relative fluorescent units

Whole-genome sequencing of the virus (named hereafter uady-tam_2025) from patient plasma resulted in 94.9% genome coverage. Comparison to the genome obtained from the viral isolate (P0) showed 99.8% nucleotide identity. Phylogenetic analysis including all publicly available full-length CHIKV genomes with complete sampling dates placed uady-tam_2025 within the ECSA lineage, clustering with genomes sampled in Brazil between January and February 2025 (SH-aLRT = 0.79) (Figure 2A, Figure S1). By contrast, all previously reported CHIKV genomes all sampled during the 2014-2015 Mexico outbreak, clustered within the Asian lineage (Caribbean, Central America and North America Clade [CCNA]) (Figure 2a) (*9,10*). Our analysis also shows synchronous circulation of multiple CHIKV lineages globally as of 2025, associated with geographically distinct outbreaks. Notably, multiple ECSA sub lineages have given rise to large ongoing outbreaks in Indian Ocean islands, southern Asia, and South America (Figure S1).

**Figure 2.**
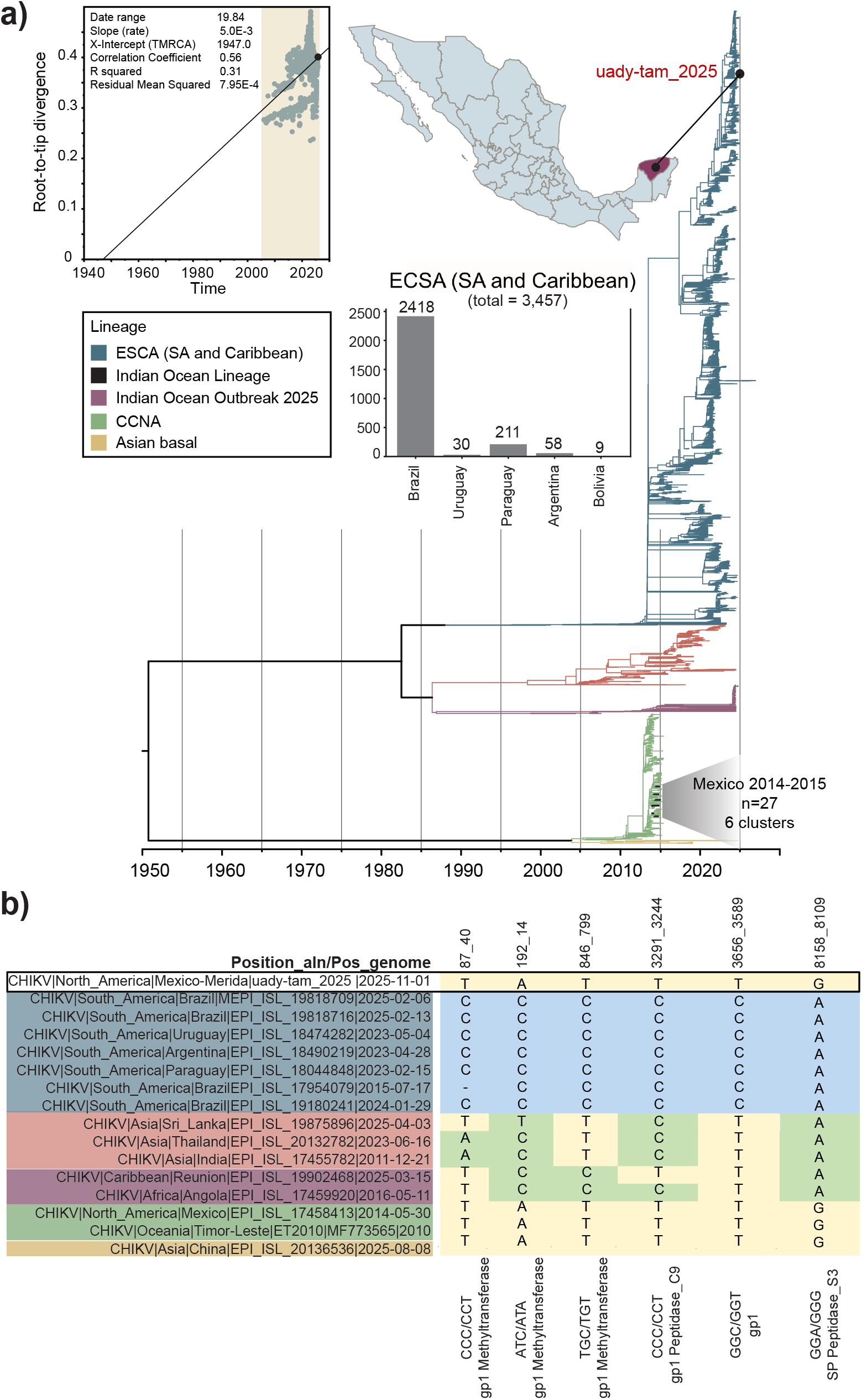
Phylogeny of CHIKV and analysis of signature synonymous mutations. (a) Maximum likelihood time-scaled phylogenetic analyses of 4651 CHIKV genomes, with branches coloured according to major CHIKV lineages and sub lineages. The uady-tam_2025 genome is indicated with a black dot on the tree within the ESCA (SA and Caribbean) lineage. The number of genomes available from this sub lineage from distinct countries in South America are indicated. Genomes sampled from Mexico during 2014-2015 CHIKV epidemic clustering within the CCNA linage are also shown. The inset displays the root-to-tip regression used to assess the temporal signal of the dataset, and the shaded region highlights the tip sampling period. **(b)** Single nucleotide polymorphisms (SNPs) representing synonymous mutations in uady-tam_2025 identified by mapping sites relative to the genome positions and coding regions of CHIKV reference genome (NC_004162). Lineage-specific SNP patterns highlighting the differences between uady-tam_2025 and contemporary ESCA genomes are shown in blue. SNP comparison with other more distantly related lineages are further highlighted in light yellow or green.

The ECSA lineage was introduced into Brazil in 2014 and has since become endemic, with diversification into distinct clades (*11*) that have been detected through genomic sequencing in several countries in South America (Figure 1a). The uady-tam_2025 genome contained the lineage defining mutations recently described in the ECSA clade II from Brazil (nsP2-P352A [C2735G], nsP4-A43V [C5793T], and E1-T288I [C10856T) (*12*). It also carried mutation E1-T98A (G10285A), associated with enhanced replication in *Ae. albopictus* within the genomic background of mutation E1-A226V, although the latter has not been detected in ECSA genomes circulating in the Americas (*13*). A long branch leading to uady-tam_2025 suggests substantial under-sampling and continued virus evolution (Figure 2a, Figure S1). In this context, the uady-tam_2025 genome contained multiple synonymous mutations absent from the parental clade (Brazil ECSA clade II) but found in other more distantly related lineages, particularly the Asian lineage. These mutations may be indicative of convergent evolution, reversions, or retention of ancestral characters (Figure 2b). In addition, non-synonymous substitutions (nsP1-R144K [G507A], nsP2-L494I [C3161A], nsP3-L434P [T5376C], nsP3-E333D [A5074T] and nsP3-D518Y [G5627T]) were also absent from the parental clade. Though their evolutionary origin cannot currently be determined, these mutations could serve as molecular markers to identify currently circulating ECSA sub lineages, as data from Cuba and/or the Caribbean is generated.

## Conclusions

Detection of CHIKF in Mexico from a traveler arriving from Cuba, a country with an active large-scale CHIKV outbreak, together with recent case reports from Yucatan and Quintana Roo, raises important public health concerns. The introduction of the ECSA lineage of CHIKV into Mexico, a lineage with evidence for heightened virulence compared to the CCNA clade (*10*), has not previously been documented. This event highlights the importance of international air travel through major tourist hubs as a pathway of viral introduction in addition to previous associations with overland migration routes through Central and North America (9). With ten years having elapsed since Mexico’s major chikungunya epidemic (2014–2015), population-level immunity is likely to have waned (14), increasing susceptibility to renewed transmission in Aedes-endemic urban settings such as the Yucatán Peninsula.

Strengthened regional collaboration for genomic monitoring will be necessary to better characterize virus circulation across Central America, North America, and the Caribbean. The current paucity of publicly available genomic data from much of the Caribbean hampers reconstruction of outbreak origins, transmission pathways and evolutionary history. Nevertheless, our phylogenetic analysis links the ongoing Cuban outbreak to the Brazil ECSA clade II (12) and, together with the placement of the uady-tam_2025 genome, suggests an expanding geographic range of this lineage into the Caribbean and potentially North America.

Travelers with high viremia returning to areas with established presence of *Ae. aegypti* and *Ae. albopictus* populations may initiate local transmission, especially in endemic regions such as the Yucatan Peninsula (*7,8,9,14,15*). Thus, public health laboratories in collaboration with local academic institutions should monitor returning travelers and conduct vector surveillance in high-risk areas to prevent establishment of emerging outbreaks. Clinicians should routinely have access to travel histories from patients presenting a CHIKF-compatible illness and should record unique symptoms of CHIKF for a differential diagnosis. Although the ecological drivers shaping the establishment of the ECSA lineage in Mexico remain under investigation, its introduction raises concern for subsequent geographic spread. Monitoring adaptive mutations—such as E1-A226V emerging within a permissive genomic background— should be integrated into ongoing genomic surveillance efforts, given that viral adaptation in local mosquito populations could enhance vector competence and facilitate sustained autochthonous transmission. Early detection of imported cases, combined with integrated clinical, entomologic and genomic surveillance as well as vector control, is critical to target potential local transmission and prevent the establishment of new epidemic CHIKV lineages in Mexico.

## Data Availability

All data produced in the present work are contained in the manuscript.

## Acknowledgements

This research was supported by the Wellcome Trust Infectious Disease Award (317324/Z/24/Z). Additionally, it was partially funded by UADY internal research project: CIRB-2025-0012. MGK is funded through a Sanger International Fellowship award. MEZ is funded by a UCL Rosetrees Excellence Fellowship UCL2024\2. We gratefully acknowledge all data contributors, i.e., the authors and their originating laboratories responsible for obtaining the specimens, and their submitting laboratories for generating the genetic sequence and metadata and sharing via the GISAID Initiative.

---

**Supplementary Figure 1.**
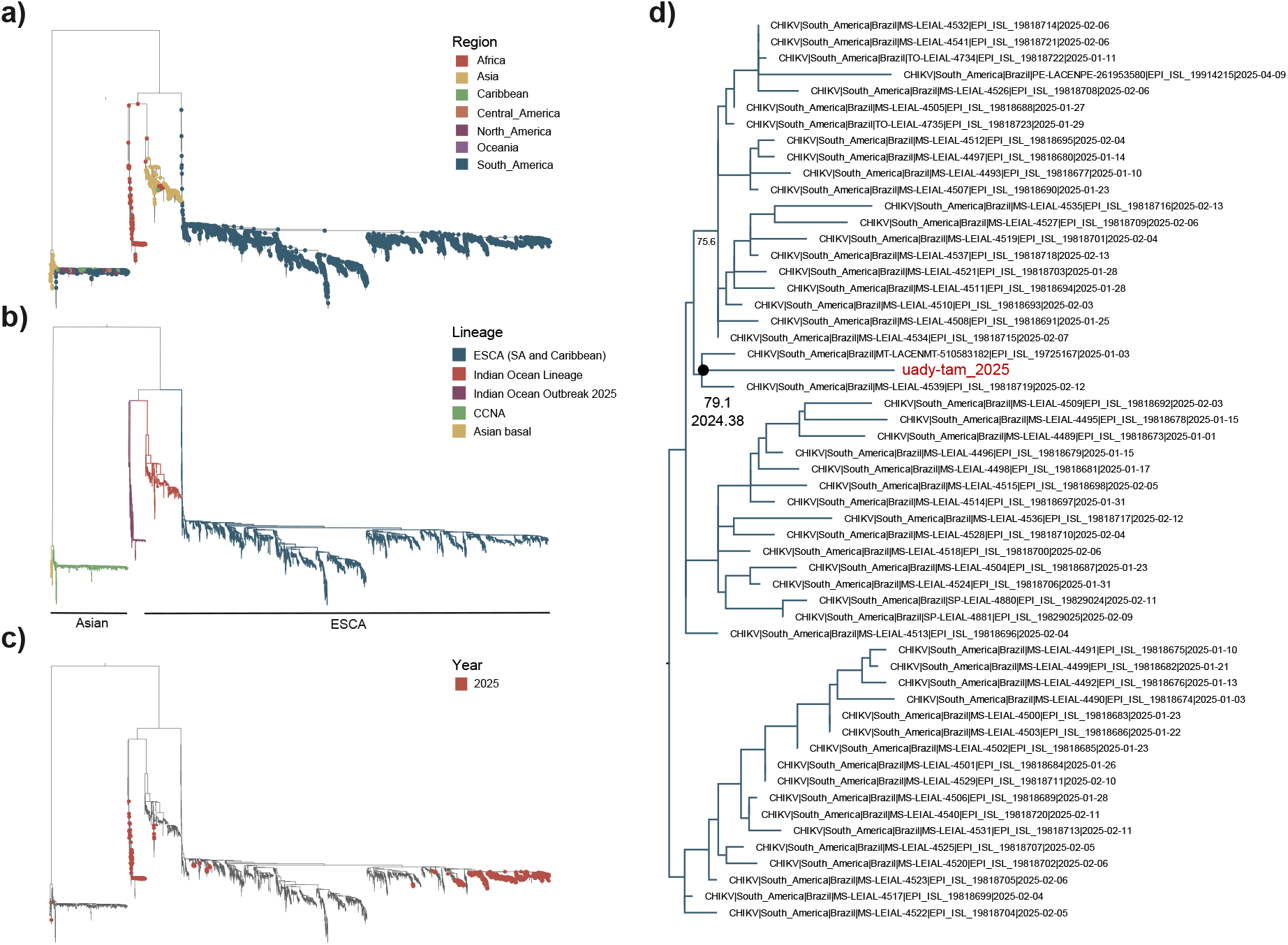
Annotated maximum-likelihood tree and high-resolution placement of uady-tam_2025 in the context of CHIKV genomes sampled globally. The ML tree is annotated by **(a)** region, **(b)** lineage, and **(c)** year of sampling (2025), whilst **(d)** a detailed zoom into the ESCA (SA and Caribbean) cluster, indicate the placement of uady-tam_2025 close to isolates sampled from Brazil between January and March 2025. A bootstrap support value (SH-aLRT) of 79.1 and maximum-likelihood point estimate for node age (2024.38) are indicated for the node/branch corresponding to uady-tam_2025. A moderate relative branch length indicates some divergence between uady-tam_2025 and the most closely related isolates identified.

