## Supplementary Materials for "TRAVEL-ASSOCIATED CHIKUNGUNYA VIRUS INFECTION, MEXICO, 2025"

**Supplemental Methods**

*RNA extraction, virus isolation and RT-qPCR for Arboviruses.*

Viral RNA was extracted from 150 μL of plasma or infected cell supernatant using the QIAamp Viral RNA Mini Kit following the manufactures instructions. Extracted RNA was quantified using a Thermo Scientific Nanodrop One Microvolume UV-Vis spectrophotometer (Thermo Scientific), aliquoted and then stored at -80°C until further analyses. A 100 μl of fresh plasma was mixed with 3 x 10^6^ Vero cells (CCL-81, ATCC) while these were still in suspension in 1 mL of DMEM media supplemented with 2% fetal bovine serum (FBS), antibiotics (Penicillin/Streptomycin) and HEPES (1X). Cell monolayers were incubated under standard cell culture conditions (5% CO2, 37C, and 80-95% relative humidity) until the cytopathic effect (CPE)was identified. Cell-free supernatants were aliquoted and stored for further analysis at -80°C. For Trioplex RT-*q*PCR, a set of DENV, ZIKV, and CHIKV primer specific primers were used as previously described (*1*). The real-time quantitative PCR (RT-qPCR) protocol was run using a Bio-Rad CFX96TM Real-Time System (Hercules, CA) as follows: 50°C for 15 minutes (one cycle) and 95°C for 5 minutes (one cycle), followed by 40 cycles of 95°C for 30 seconds and 57.5°C for 30 seconds. Data analyses were performed using the Bio-Rad CFX master software v 2.3. Human samples were considered positive if cycle threshold (CT) values were below 35. RNA extracted from DENV, ZIKV, and CHIKV prototype strains were used as a positive controls as following described: DENV (pool RNA from serotype 1, 2, 3 and 4 RNA): Ct 36; ZIKV (MR-766 strain): Ct 26.44; CHIKV (181/25): 24.99. DENV strains included in the RNA pool: DENV-1 (West Pac 74); DENV-2 (S-16803); DENV-3 (CH53489); DENV-4 (TVP-360).

Written informed consent was obtained from the patient. Data was anonymized, and nonessential identifying details were omitted to protect patient privacy. This report is based on the regulations applicable in Mexico, such as the General Health Law and the Official Mexican Standard NOM-012-SSA3-2012 on health research. It focuses on presenting laboratory results from a specific case as a descriptive analysis, which did not involve invasive procedures or significant risks to the patient; therefore, it was exempted from formal evaluation by the ethics committee of UADY.

*Genome sequencing and assembly.*

RNA extracted from patient plasma or virus-infected cell supernatants was converted to cDNA using LunaScript RT Super Mix (New England Biolabs). Tiling-amplicons spanning the viral genome were generated by multiplex PCR using published primers sets and protocols (*2*). The PCR product yield was evaluated via agarose gel (1.5%) electrophoresis separation, and visualization by staining with SYBR™ Green I and ChemiDoc™ MP System with Image Lab™ Software. DNA quantification was done on the Qubit 3.0 instrument (Life Technologies) using the Qubit dsDNA High Sensitivity assay. Libraries were synthesized with the ligation sequencing DNA kit (SQK-LSK114.24, Oxford Nanopore Technologies) and samples were barcoded using the Native Barcoding Kit (NBD104, Oxford Nanopore Technologies, Oxford, UK). Sequenced was done for 72 hours on a MinION instrument (Oxford Nanopore Technologies) using R10.4.1 flow cells. Raw FATS5 files were base called and demultiplexed with Guppy v6.5.7 (https://github.com/nanoporetech). The Genomic Detective Platform was used for primer trimming, read alignment, variant calling and consensus assembly (*3*).

*Sequence dataset and alignment*

The global dataset of complete, high coverage CHIKV genomes (n = 4,888) sampled between 2005 and 2025 was downloaded from the GISAID database (*4,5,6*) available as of 27th November 2025 via *https://doi.org/10.55876/gis8.251208mo*. Genomes from Mexico reported by Gutierrez et al. 2023 were also included (*7,8*). Sequences were aligned using MAFFT v7 (*9*) under default parameters. The alignment was manually inspected to remove misaligned regions and low-quality sequences. Identical sequences were pruned prior to phylogenetic analysis. After removing sequences belonging to the phylogenetically distant West African lineage and outliers identified via root-to-tip regression using TempEst (*10*), the final dataset comprised 4,651 sequences.

*Maximum-likelihood phylogenetic reconstruction*

A maximum-likelihood (ML) tree was reconstructed using IQ-TREE 2 v2.2 (*11*) under a GTR+G substitution model and SH-aLRT branch support (1,000 replicates). Information on sampling region/country and date were extracted from the sequence metadata to annotate the ML tree using FigTree v1.4.4 (https://tree.bio.ed.ac.uk/software/figtree/), whilst CHIKV lineages were annotated in reference to described most recent phylogenetic analyses (*7,12,13*).

*Time-calibrated phylogeny*

A time-scaled phylogeny was reconstructed from the ML tree using TreeTime v0.9.5 (*14*) inputting sampling dates extracted from the sequence metadata. A second run incorporated a fixed clock rate (1.5x10^3^ subs/site/year) consistent with reported CHIKV evolutionary rates (*12*).

*Mutation characterization*

Single nucleotide differences between the uady-tam_2025 isolate and those belonging to identified CHIKV lineages (including ESCA) were identified through pairwise sequence comparison relative to the earliest and latest sampled sequence belonging to each lineage. Single nucleotide polymorphisms (SNPs) were annotated, with the corresponding synonymous/non-synonymous mutations identified based on the CHIKV reference genome annotations (NC_004162) and confirmed in NextClade (https://clades.nextstrain.org) and the ChikSurver tool in GISAID (https://gisaid.org/).

**References (Supplemental Material)**
